# Prevalence clinical correlates and outcomes of cardiorenal anemia syndrome among patients with heart failure attended a tertiary hospital in Dodoma, Tanzania: A prospective observational cohort study

**DOI:** 10.1101/2024.06.13.24308917

**Authors:** Gidion Edwin, Baraka Alphonce, Alfred Meremo, John Robson Meda

## Abstract

**Introduction:** Cardiorenal anemia syndrome (CRAS) poses significant complications in heart failure (HF) patients, often leading to unfavourable outcomes but, published data are limited. This study assessed the prevalence, clinical correlates, and outcomes of CRAS among patients with HF who attended the Benjamin Mkapa Hospital (BMH) in Dodoma, Tanzania.

**Materials and methods:** A prospective observational cohort study was carried out at BMH between 18^th^ August 2023 and 18^th^ April 2024. It included patients aged 18 years and above who had been diagnosed with heart failure according to Framingham criteria and confirmed by 2-dimensional transthoracic echocardiography (2D-TTE). The study aimed to investigate the prevalence and clinical associations of cardiorenal anemia syndrome (CRAS) at the beginning of the study, as well as to evaluate CRAS outcomes within a 6-month follow-up period. Continuous data were presented as either mean with standard deviation (SD) or median with interquartile range (IQR), while categorical data were expressed as frequency and proportions. Binary logistic regression, using odds ratios (OR), was utilized to examine clinical associations, while survival rate analysis, employing hazard ratios (HR), was utilized to determine CRAS outcomes. A two-tailed p-value of less than 0.05 was considered statistically significant.

**Results:** A total of 298 participants were recruited with a mean age of 57±15 years, and 60% were females. In our cohort, CRAS was prevalent in 46.3%. Iron deficiency (OR: 2.5; 95% CI, 1.5-4.1; *p* = 0.001) and diabetes mellitus (OR 2.1; 95% CI, 1.2-3.4; *p* = 0.006), were clinically correlated with CRAS, while female sex (OR 0.35; 95% CI, 0.21-0.59; *p* = 0.000) was inversely clinically correlated with CRAS. Moreover, CRAS was associated with a higher risk of heart failure re-hospitalization compared to those patients with no CRAS (HR: 3.8; 95% CI, 2.4-6.0; *p* < 0.001).

**Conclusion:** In our setting, CRAS is prevalent among heart failure patients and is linked to higher rates of heart failure-related hospitalizations, leading to increased healthcare utilization and costs. We strongly advocate for multidisciplinary approaches in managing this condition. Nonetheless, further research with robust evidence is necessary to inform policy-making and initiate targeted interventions.

## Introduction

Cardiorenal anemia syndrome (CRAS) is a complex condition that includes heart failure of any type, chronic kidney disease with an estimated glomerular filtration rate of less than 60 ml/min/1.73 m², and anemia with haemoglobin levels below 13 g/dl for men or 12 g/dl for women (1). CRAS poses a significant global public health challenge, particularly among heart failure patients, as it is linked to poor clinical outcomes (2,3).

The prevalence of CRAS is widely recognized ranging from 4.6% to 35.4% in developed countries (1,2,4) and from 19% to 62% in developing countries (5,6). Previous studies have shown that, advanced New York Heart Association (NYHA) class, diabetes mellitus, iron deficiency, and gender were clinically associated with CRAS portending an unfavourable prognosis (1,3,6). Follow-up studies investigating CRAS outcomes among heart failure patients with reduced ejection fraction (HFrEF) demonstrated its association with elevated risk of all-cause mortality, cardiovascular mortality, heart failure rehospitalization, and deterioration in renal function. These outcomes were attributed to increased healthcare costs and utilization (7–11).

In Tanzania, CRAS was found in 44.4% of patients with HFrEF, and it was linked to a higher risk of all-cause mortality and heart failure hospitalization during follow-up (6). However, there is currently insufficient data on CRAS across all heart failure phenotypes in our context. This study seeks to fill this gap by investigating the prevalence, clinical associations, and outcomes of CRAS among patients at Benjamin Mkapa Hospital in Dodoma, Tanzania, across various HF phenotypes.

## Materials and methods

### Study design and settings

This prospective observational cohort study was conducted at Benjamin Mkapa Hospital (BMH) in Dodoma, Tanzania, a public tertiary hospital located within the University of Dodoma (UDOM) campus, 13 kilometres from Dodoma city centre. BMH is a specialized tertiary hospital with comprehensive cardiology services, including cardiac imaging and interventional cardiology. The hospital has a capacity of 400 beds and serves approximately 20 to 50 heart failure patients daily in its clinic. Additionally, BMH functions as a teaching hospital for both undergraduate and graduate programs at the University of Dodoma.

### Study participants

Participants (human subjects) were patients with heart failure aged 18 years or older who consented by signing the written informed consent form were recruited in this study. No minors were enrolled.

### Sample size and sampling method

The estimated sample size for this study was determined using a method for proportion in a prospective observational cohort study, based on a previous study conducted in Tanzania(12,13). A minimum of 160 participants was required to meet the expected sample size. However, to enhance the study’s power and effect size, 298 participants were enrolled through convenient serial sampling over nine months from 18^th^ August 2023 to 18^th^ April 2024. Recruitment period started from 18^th^ August, 2023 to 18^th^ November, 2023 and then, all participants were followed for 6 months on monthly basis from day of recruitment until 18^th^ April, 2024.

### Inclusion and exclusion criteria

Only consenting adult patients aged 18 years or older, diagnosed with heart failure according to the Framingham criteria and confirmed by 2-D TTE, with chronic kidney disease (estimated GFR < 60 ml/min/1.73 m²) and anemia (haemoglobin levels < 13 g/dl for males or < 12 g/dl for females), were consecutively enrolled in the study. Exclusions included patients with known solid or haematological malignancies undergoing treatment, pregnant patients, those with obvious gastrointestinal bleeding, known Hemoglobinopathies, patients on iron supplementation, those with end-stage renal disease (ESRD) on dialysis, and patients who had received a pacemaker or implantable cardioverter defibrillator.

### Outcomes variables

#### Primary outcome

The primary outcome of this study was cardiorenal anemia syndrome (CRAS), which was assessed as a combination of heart failure (regardless of ejection fraction), chronic kidney disease (estimated glomerular filtration rate <60 ml/min/1.73 m²), and anemia (haemoglobin levels < 12 g/dl for women or < 13 g/dl for men) (1,6)

#### Secondary outcomes

Secondary outcomes included:

**All-cause mortality** was defined as any death that occurred within 6 months of follow-up irrespective of cause (1,6,14,15)

**Heart failure rehospitalization** was defined as having more than one admission related primarily to heart failure (4,6,10,14,16)

### Independent variables

#### Participant’s enrolment

Data collection was conducted using a well-structured questionnaire. From 18^th^ August 2023 to 18^th^ April 2024 (recruitment period from 18^th^ August, 2023 to 18^th^ November, 2023); a total of 298 patients who met the inclusion criteria were enrolled. They were followed for 6 months to observe secondary outcomes. Follow-up was conducted monthly via phone calls or direct contact during clinic visits on monthly basis. At the beginning of the study, socio-demographic data such as age, sex, and cigarette smoking status were collected. For anthropometric measurements, waist and hip circumferences were recorded, and the waist-hip ratio was calculated. (17). Furthermore, information regarding NYHA functional class, diabetes mellitus, hypertension, dyslipidaemia, obesity, anemia, chronic kidney disease, and iron deficiency was collected at baseline.

**Waist-hip ratio (WHR)**: The Waist-Hip Ratio (WHR) was defined as the ratio of waist circumference to hip circumference. A WHR of ≥ 0.9 for men or ≥ 0.8 for women indicates central obesity. Waist circumference was measured with the individual standing upright, abdomen relaxed, arms at the sides, and recorded at the smallest diameter between the rib cage and the pelvic bone, with central obesity defined as > 102 cm for men and > 88 cm for women. Additionally, hip circumference in centimetres was measured over minimal clothing at the level of the greater trochanter, around the widest part of the buttocks.

**Blood pressure (BP) measurements:** BP was measured using a calibrated automated BP machine, following the conventional method outlined in the 2018 AHA/ACC hypertension guidelines. A blood pressure reading of ≥ 140/90 mmHg or the use of anti-hypertensive medication indicated hypertension. (6,12,18).

#### Laboratory investigations

All blood samples were collected according to the laboratory standard operating procedures accredited and supervised by Benjamin Mkapa Hospital. The samples were analysed using the MAGLUMI 800 CLIA (China, 2019) and Roche Cobas 6000 CLIA (USA, 2018) clinical chemistry automatic analysers. The following investigations were performed:

**Lipid profile measurements:** Dyslipidaemia was defined as having any of the following: total serum cholesterol ≥ 200 mg/dL, low-density lipoprotein cholesterol (LDL-C) ≥ 130 mg/dL, serum triglycerides ≥ 150 mg/dL, high-density lipoprotein cholesterol (HDL-C) < 40 mg/dL for women and < 50 mg/dL for men, or the use of lipid-lowering medications. (12,19,20)

**Blood Sugar measurements:** Diabetes mellitus (DM) was defined as having glycated haemoglobin (HbA_1_C) ≥ 6.5% or evidence of using anti-diabetic medications. HbA_1_C provides greater accuracy in diagnosing DM (21–23)

**Full blood picture:** Anemia was defined as haemoglobin < 12 g/dl for females or < 13g/dl for males. (6,12,24,25).

**Iron studies:** Iron deficiency was defined as having serum ferritin levels < 100 µg/L, or serum ferritin levels between 100 and 300 µg/L with a transferrin saturation (TSAT) ≤ 20%. (24,26–29).

**Serum creatinine measurement**: Chronic kidney disease was identified using the estimated glomerular filtration rate (eGFR) calculated with the CKD-EPI formula, adjusted for the African-American correction factor. An eGFR value of < 60 ml/min/1.73 m² was used to define chronic kidney disease (22,30,31).

#### Cardiac imaging

All patients underwent 2-dimensional transthoracic echocardiography (model Vivid TM T9, manufactured by GE Healthcare, USA, 2018). The ejection fraction and diastolic or systolic dysfunction were used to classify heart failure phenotypes (HFpEF, ≥ 50%; HFrEF, < 50%) (32). Two consultant cardiologists interpreted and confirmed all echocardiography reports.

### Follow-up procedure

Each patient received monthly follow-ups for 6 months. Throughout this timeframe, patients were monitored for specific outcomes, including worsening heart failure symptoms, heart failure rehospitalization and all-cause mortality. The dates when these outcomes occurred were documented.

### Data analysis

During statistical analysis, the data were entered into a Microsoft Excel spreadsheet and then transferred into the SPSS Windows version 26 program (IBM SPSS, Chicago IL) (12). Data were classified as CRAS or non-CRAS groups at baseline. Data were reported as mean with standard deviation (S.D) or median with interquartile range (IQR) for continuous variables while frequencies and proportions were used for categorical variables. The clinical correlates were assessed by binary univariate logistic regression, and the only variables that met at least a 20% (*p*-value ≤ 0.2) statistical significance (3,12,14), were selected for multivariable logistic regression. The Cox proportional hazards model survival analysis were used to evaluate the association between CRAS and 6-month outcomes. The hazard ratio and 95% Confidence interval were shown. A *p*-value of less than 0.05 was considered statistically significant (1,4).

### Ethical issues

This study adhered to the ethical principles outlined in the Declaration of Helsinki. Ethical clearance was obtained from the Directorate of Research and Publications (reference number: **MA.84/261/64/134**), and authorization for the study was granted by the Vice Chancellor’s office at the University of Dodoma. A permit letter for data collection was provided by the administrative division of Benjamin Mkapa Hospital in Dodoma (reference number: **AB.150/293/01/” A”/34**). Participants were informed by signing the “written informed consent form” that their involvement was voluntary, and they had the right to withdraw at any time. To protect privacy and confidentiality, participants’ identities were replaced with identification numbers. It’s important to note that their decision to participate did not affect the standard of care they received.

## Results

Out of 298 participants who met the inclusion criteria, the prevalence of CRAS was 46.3%. Finally, we had 268 patients who were analyzed for outcomes of interest.

**Fig. 1.**
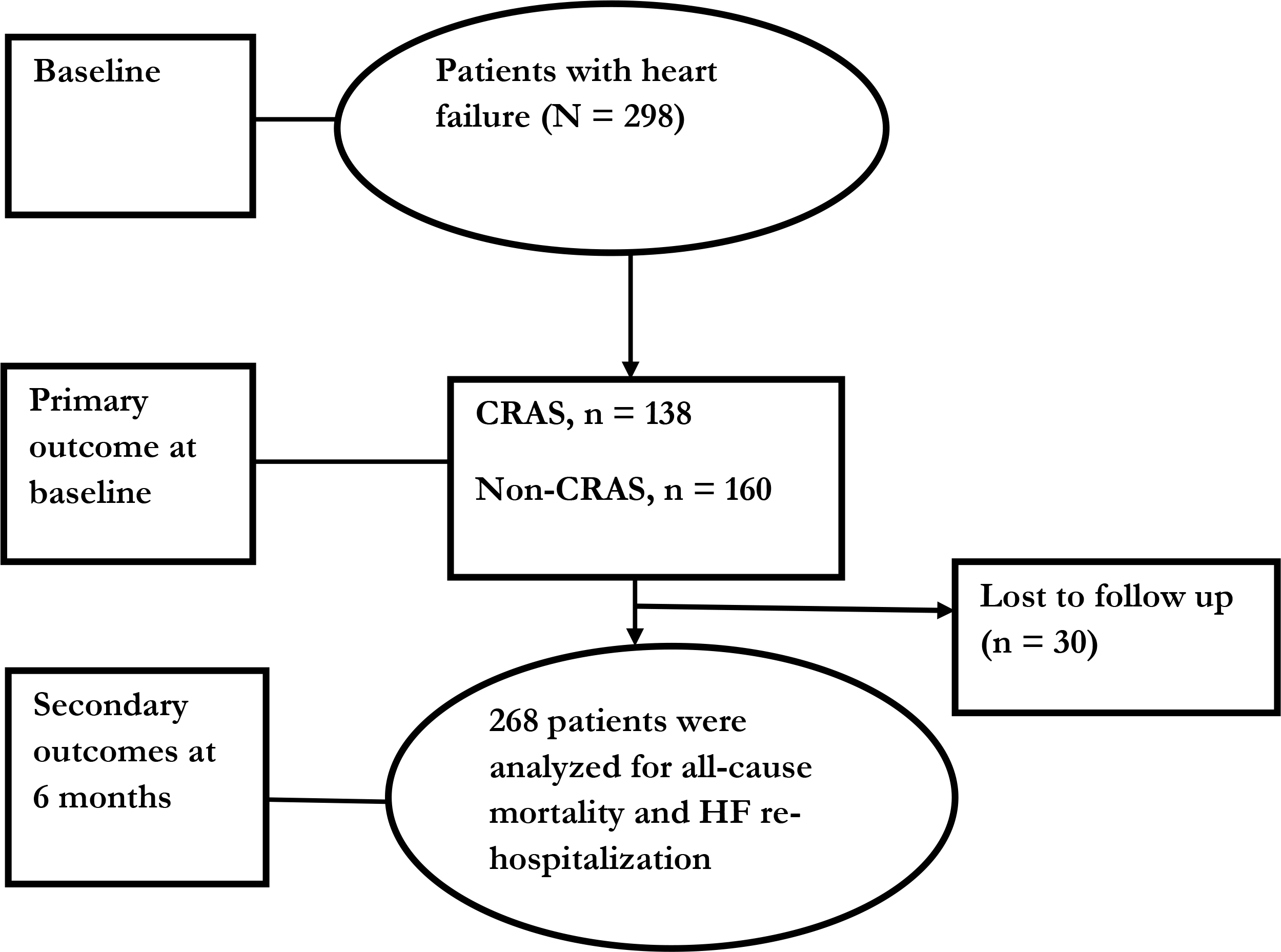
Algorith1n flowchart of the study populations by CRAS.

### Socio-demographic characteristics

The mean age of the participants in the study was 57±15 years. Women constituted 60% of the study population (Table 1). The majority of participants 72% (213) were below the age of 65. At baseline, the prevalence of hypertension was 75%, diabetes affected 51% of participants, while 46% had dyslipidaemia 60% of participants were current smokers, and 48% were classified as obese.

**Table 1.** Baseline characteristics of the study participants.

| **Variables** | **Category** | **All**<br>**(N = 298)** | **No CRAS**<br>**(n = 160)** | **CRAS**<br>**(138)** | ***p*-value** |
| --- | --- | --- | --- | --- | --- |
| **Age, in years** | Mean (S.D) | 57 (15) | 55 (14) | 58 (15) | 0.058 |
| **Age group**<br>**(years)** | < 65 | 213 (72) | 117 (73) | 96 (70) | 0.291 |
|  | ≥ 65 | 85 (28) | 43 (27) | 42 (30) |  |
| **Sex** | Female | 178 (60) | 76 (48) | 102 (74) | 0.001 |
|  | Male | 120 (40) | 84 (52) | 36 (26) |  |
| **Smoking** | Yes | 118 (60) | 71 (44) | 47 (34) | 0.229 |
|  | No | 180 (40) | 89 (56) | 91 (66) |  |
| **Obesity (waist-**<br>**hip ratio)** | Yes | 142 (48) | 84 (53) | 58 (42) | 0.04 |
|  | No | 156(60) | 76 (47) | 80 (58) |  |
| **Hypertension** | Yes | 224 (75) | 117 (73) | 107 (78) | 0.38 |
|  | No | 74 (25) | 43 (27) | 31 (22) |  |
| **Diabetes**<br>**mellitus** | Yes | 153 (51) | 73 (46) | 80 (58) | 0.02 |
|  | No | 145 (49) | 87 (54) | 58 (42) |  |
| **Dyslipidaemia** | Yes | 136(46) | 68 (42) | 68 (49) | 0.246 |
|  | No | 162(54) | 92 (58) | 70 (51) |  |
| **Anemia** | No | 128 (48) | 128 (89) | 0 (0) | 0.001 |
|  | Yes | 140 (52) | 16 (11) | 125 (100) |  |
| **CKD** | No | 67 (25) | 67 (46) | 0 (0) | 0.001 |
|  | Yes | 201 (75) | 76 (54) | 125 (100) |  |
| **Iron deficiency** | No | 109 (68) | 67 (48) | 176 (59) | 0.001 |
|  | Yes | 51 (32) | 71 (52) | 122 (41) |  |
| **NYHA functional class** | I | 13 (4) | 6 (4) | 7 (5) | 0.001 |
|  | II | 119 (40) | 81 (51) | 38 (27) |  |
|  | III | 100 (34) | 47 (30) | 53 (38) |  |
|  | IV | 66 (22) | 26 (15) | 40 (30) |  |
| **Heart failure phenotypes** | HFrEF | 158 (53) | 75 (47) | 83 (60) | 0.015 |
|  | HFpEF | 140 (47) | 85 (53) | 55 (40) |  |

### Clinical correlates of CRAS

Under adjusted multivariable logistic regression analysis, iron deficiency (OR 2.5; 95% CI, 1.5-4.1; *p* = 0.001); diabetes mellitus (OR 2.1; 95% CI, 1.2-3.4; *p* = 0.006), and female sex (OR 0.35; 95% CI, 0.21-0.59; *p* < 0.001) were identified as significant clinical correlates of CRAS (**Error! Reference source not found.**).

**Table 2.**
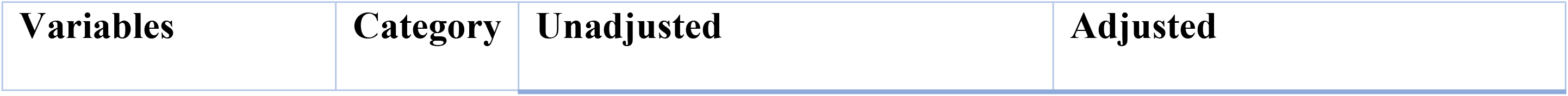
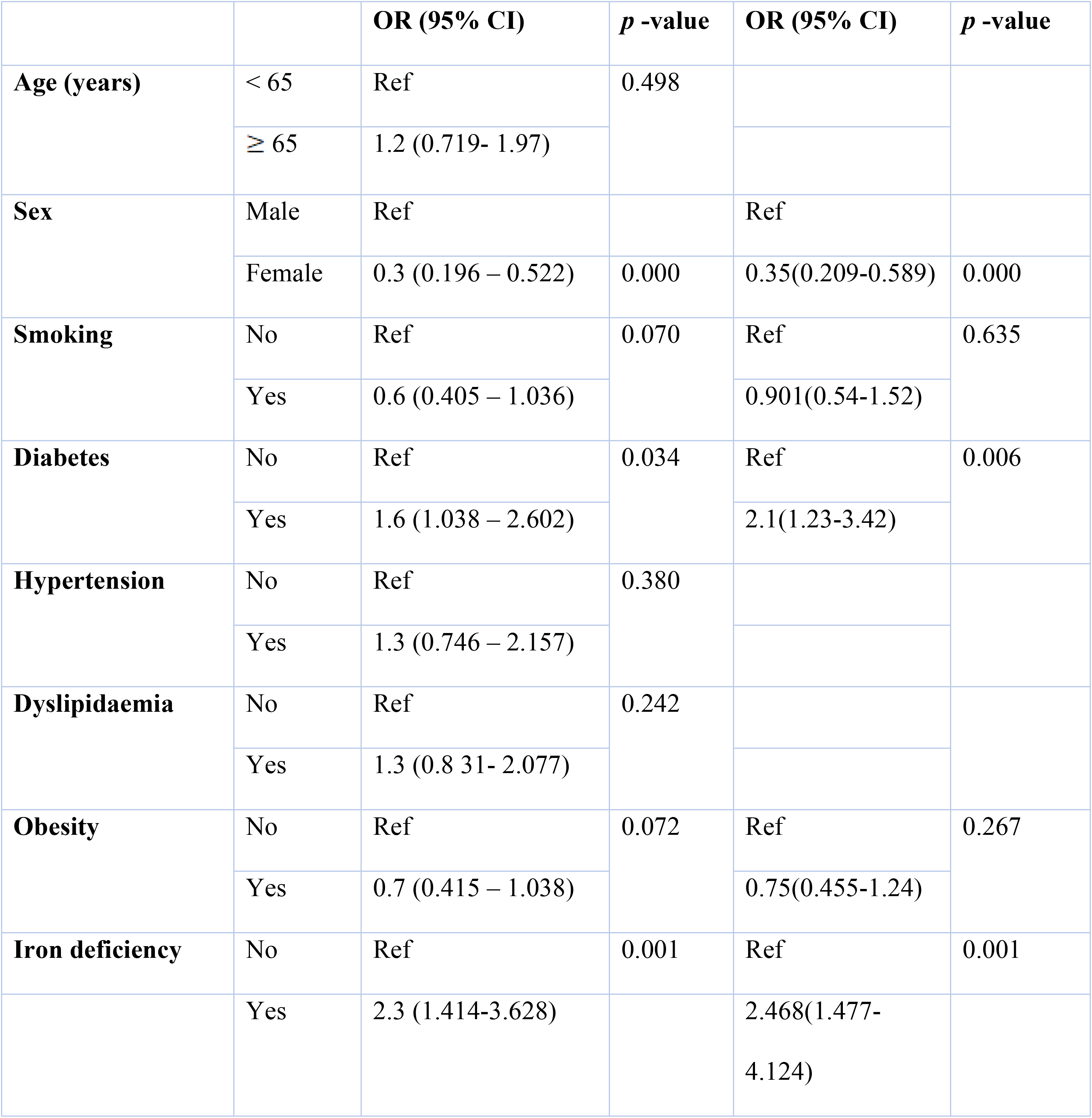
Clinical correlates of CRAS.

### Outcomes of CRAS

Overall, patient with CRAS demonstrated 3.8 times risk for heart failure rehospitalisation rate compared to non – CRAS patients (HR 3.8, 95% CI 2.4 – 3.6, *p* < 0.001) (**Error! Reference source not found.** and **Error! Reference source not found.**). However, there was no difference in risk of all-cause mortality between the CRAS groups (HR 1.4, 95%CI 0.9, *p* = 0.101).

**Fig 2.**
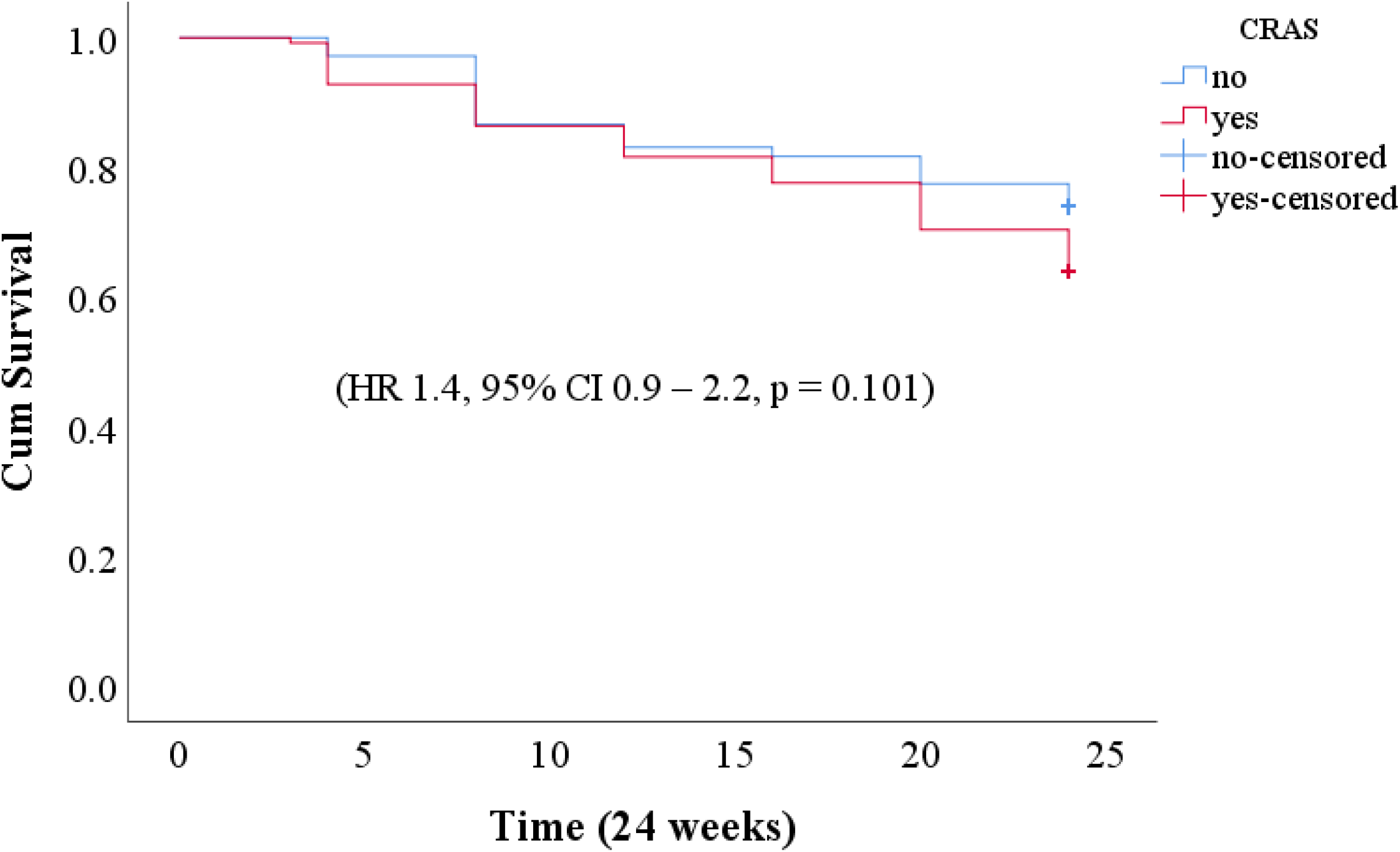
Cox proportional hazard model survival curve showing All-cause mortality by CRAS.

**Fig. 3.**
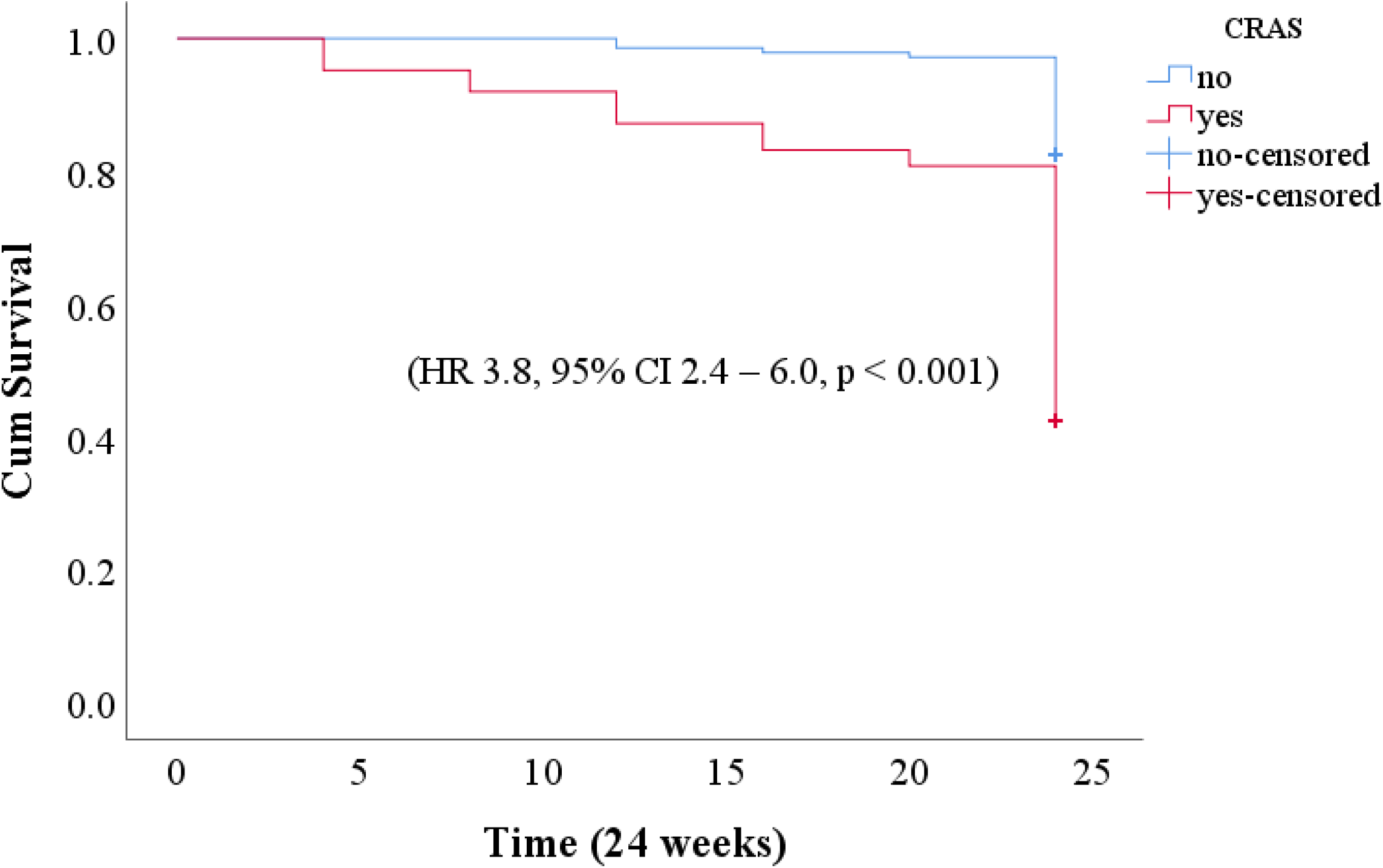
Cox proportional hazard model survival curve showing HF rehospitalization by CRAS.

## Discussion

The primary aim of this study was to assess the prevalence, clinical associations, and outcomes in patients with heart failure across various phenotypes at Benjamin Mkapa Hospital in Dodoma, Tanzania.

In our study group, the prevalence CRAS across HF phenotypes was 46.3%. This aligns closely with the findings of Belmar Vega et al., who reported a CRAS prevalence of 46% in their cohort across HF phenotypes (5). Similarly, an earlier study conducted in Tanzania focusing on patients with HFrEF reported a prevalence of 44.4% (6). Our results are consistent with previous research, which has shown CRAS prevalence rates ranging from 19% to 62% among the patients with HF across phenotypes (33–35).

Previous studies showed that the presence of comorbid conditions such as diabetes, hypertension, and chronic kidney disease and iron deficiency with or without anemia predicted the occurrence of CRAS. In such context, the similarity in CRAS prevalence across previous studies and our cohort could be explained by sharing of common determinants.

We found that CRAS correlated clinically with male sex, iron deficiency, and diabetes mellitus. The finding aligns with previous studies that have highlighted the role of iron metabolism in the pathophysiology of CRAS (2,20,36–38). Scrutinio et al., demonstrated that iron deficiency, irrespective of the presence of anemia, significantly impacts CRAS outcomes by exacerbating heart and kidney dysfunctions (1). The chronic inflammatory state in heart failure patients leads to impaired iron absorption and utilization, further portending the clinical severity of CRAS (39).

In the Middle East, a previous study found that males were more often linked to CRAS than females. This was partly due to the prevalence of comorbid conditions such as diabetes and hypertension among males. Additionally, unequal access to healthcare services may have also contributed to this disparity (14). In Sub-Saharan Africa including our setting, poor accessibility to health care services and the presence of underlying risk factors such as cigarette smoking might contributed the effect of the male sex towards the hypothetical pathogenic domains of CRAS (6). In our cohort, females were inversely clinically correlated with CRAS and hence, males were found to be clinically correlated with the occurrence with CRAS; similar across previous studies globally (6,14).

Studies from the Middle East, Japan, and Tanzania has demonstrated that diabetes mellitus raises the risk of CRAS and results in a poorer prognosis (6,14,40,41). Our findings also support the link between CRAS and diabetes mellitus, aligning with these earlier studies. The suggested pathogenic mechanisms included Hyperglycaemia and insulin resistance contributed to the likelihood of renal dysfunction and anemia through oxidative-reductive pathways, chronic inflammatory response, and altered erythropoiesis (39).

The former studies demonstrated that CRAS was associated with increased risk for all-cause mortality and HF-related rehospitalization (6,25,41,42). Our cohort showed HF-rehospitalization rates among patients who had CRAS were 3-fold higher than non-CRAS. Previous studies by Scrutinio et al., and Pallangyo et al., produced similar results. However, Scrutinio et al., included older patients with HFrEF, while Pallangyo et al., studied younger patients with HFrEF (1,6). CRAS accelerates structural and hemodynamic changes leading to deterioration in left ventricular and intrinsic renal functions, and therefore, portend increased rehospitalization rate and hence, worse prognosis (1,25,33).

While our study did not find a statistically significant difference in all-cause mortality between CRAS and non-CRAS patients, the trend towards higher mortality in the CRAS group is alarming. In Iraq, Goran et al. showed no statistical differences for all-cause mortality among patients with HF across phenotypes (43). Our findings are also consistent with the previous study in Brazil and Netherland among patients with HFrEF which showed no mortality difference (35). Conversely, other studies have indicated an increased risk of all-cause mortality in patients with CRAS (1,6,14,39). The lack of significant mortality difference in our cohort could be due to the relatively short follow-up period or differences in the baseline characteristics of the study population as compared to other previous studies (10,14). The majority of our study cohort were younger out-patients as compared to elderly admitted patients from developed countries. But also, could be explained by censoring the patients after the occurrence of secondary outcomes and heterogeneity of population; in our cohort, the majority were young and hence the access to health care services and adherence to medications was favourable. Nonetheless, our study cohort were seen as out-patients with chronic heart failure as compared to hospitalized patients from other studies (6,14); so, differences in interventional approaches could be hypothetical pathogenic domains toward similarity between CRAS and non-CRAS group in our findings.

## Limitations of the study

Our study has several limitations. First, the single-centered study with a minimal sample size may hinder the generalizability of our findings. Second, censoring the patients after the occurrence of secondary outcomes may not capture long-term outcomes such as mortality.

## Conclusion

In our setting, CRAS is prevalent among heart failure patients and is linked to higher rates of heart failure-related hospitalizations, leading to increased healthcare utilization and costs. Recognizing the significance of CRAS in Dodoma, we strongly advocate for multidisciplinary approaches to managing this condition. Nonetheless, further research with robust evidence is necessary to inform policy-making and initiate targeted interventions.

## Data Availability

All relevant data are within the manuscript and its supporting information files.

## Acknowledgments

The authors appreciated the patients and staffs of the Benjamin Mkapa Hospital in Dodoma for their readiness and voluntarily active participation in this study.

## Author contributions

**Conceptualization:** Gidion Edwin, Alfred Meremo, John Meda

**Data clearance and curation**: Gidion Edwin

**Formal analysis**: Gidion Edwin, Baraka Alphonce

**Investigation:** Alfred Meremo, John Meda

**Methodology**: Gidion Edwin, Baraka Alphonce, Alfred Meremo, John Meda

**Supervision**: Baraka Alphonce, Alfred Meremo, John Meda

**Writing – original draft**: Gidion Edwin

**Writing – review & editing:** Baraka Alphonce, Alfred Meremo, John Meda

